# No Point Beating Around the Bedpan: Lessons from a Major Intra-Hospital NDM-Producing *Escherichia coli* Carriage Outbreak – a Mixed-Methods Study

**DOI:** 10.64898/2026.07.06.26354129

**Authors:** Anne Le Hir, Vincent Prunet, Feyrouz Sonia Sardi, Chantal Giglione, Numida Bouton, Chloé Stavris, Lucas Maisonobe, Laurent Chiche, Carole Fliniaux, Laurent Dortet, Matthias Castanier, Jihane Brisson, Stanislas Rebaudet

## Abstract

Antimicrobial resistance constitutes a major threat to global public health. Among emerging extensively drug-resistant bacteria (eXDR), carbapenemase-producing Enterobacterales (CPE) expose hospitals to outbreaks through rapid dissemination, and to therapeutic limitations. Through a mixed epidemiological–qualitative methods study, we report the most extensive CPE carriage outbreak known to date in France, which occurred at Hôpital Européen Marseille (HEM) between January and June 2025. By the end of November 2024, the admission of an index patient returning from Senegal carrying an NDM-producing *Escherichia coli* led to an extensive transmission, despite adherence to national “screen and isolate” guidelines. Nearly 7,500 rectal screening tests identified 481 CPE carriers (including 343 NDM, 129 OXA-48-like and 9 other CPE), and 14 vancomycin-resistant *Enterococcus faecium* carriers. This major outbreak prompted an extraordinary response involving clinical, technical and administrative teams within the institution. It highlighted operational limitations in current screening, cohorting and biocleaning strategies in the context of a hospital-wide outbreak. We describe the outbreak trajectory, the control measures implemented and provide a structured synthesis of lessons learned across organisational, scientific and policy domains.

## 1 Introduction

Antimicrobial resistance (AMR) is considered one of the major health crises of the 21st century, as a threat to permanently undermine advances in modern medicine according to the World Health Organization [1]. By 2050, in the absence of drastic prevention measures, 40 million deaths related to AMR are estimated at risk to occur, especially among the rising population of people over 70 years old [1,2]. Analyses of this situation incriminate inappropriate and excessive use of antibiotics in human and veterinary medicines, particularly through uncontrolled access to antimicrobials in many countries, and the circulation of counterfeit or sub-standard drugs. AMR is present worldwide and is a major cause of concern in certain regions, such as Asia, the Mediterranean basin, and Sub-Saharan Africa, where highly resistant strains have become endemic [1,2]. Among multidrug-resistant organisms (MDRO), infections caused by extended-spectrum β-lactamase (ESBL)-producing Enterobacterales, methicillin-resistant *Staphylococcus aureus* (MRSA), carbapenem-resistant *Pseudomonas aeruginosa* (CRPA), imipenem-resistant *Acinetobacter baumannii* (IRAB), or glycopeptide-intermediate *Staphylococcus aureus* (GISA) are associated with increased morbidity and mortality [1,2]. Beyond these MDRO, there is growing concern about emerging extensively drug-resistant bacteria (eXDR), particularly carbapenemase-producing Enterobacterales (CPE) – comprising among other resistance genes *bla*_KPC_ (*Klebsiella pneumoniae* carbapenemase), *bla*_NDM_ (New Delhi metallo-β-lactamase) or *bla*_OXA-48-like_ (such as *bla*_OXA-48_, *bla*_OXA-181_, or *bla*_OXA-244_) – and vancomycin-resistant *Enterococcus faecium* (VRE) [2,3]. These organisms, part of the digestive flora, often carry resistance on mobile genetic elements, thus facilitating horizontal transfer, and spreading to new hosts both in hospital and community settings [4].

European hospitals are facing an increasing prevalence of eXDR, especially in South-Eastern Europe. According to the European Centre for Disease Prevention and Control (ECDC), 60 % of *Klebsiella pneumoniae* isolates in Greece were carbapenem-resistant in 2024, compared to 24 % in Italy and 1 % in France [5]. In Greece, this resistance is predominantly associated with *bla*_KPC-2_ and *bla*_NDM-1_ [6]. Carbapenem-resistant *Escherichia coli* remains rare (1.2 % in Greece), yet rectal carriage and infections with *bla*_NDM-5_-carrying *E. coli* have increased in the past decade, especially in France [7]. This trend is driven by intensified international travelling and excessive antibiotic pressure within healthcare systems, leading to asymptomatic intestinal carriers. Many countries such as France have responded to this issue with the establishment of “screen and isolate” guidelines [8,9], based on three pillars (Table 1): (i) systematic screening for rectal carriage of eXDR upon admission in at-risk patients, particularly those previously hospitalised abroad; (ii) identification, risk stratification and screening of contacts; and (iii) isolation and cohorting of carriers and high-risk contacts under additional contact precautions (ACP) within dedicated units, placement of newly admitted patients in separate clean zones, and adherence to the “forward progression” policy [10]. These guidelines aim to prevent secondary cases of intestinal carriage, and ultimately to avoid difficult-to-treat invasive infections. Nevertheless, they are complex to implement, thus heterogeneously applied [11,12], and sporadic nosocomial outbreaks of eXDR rectal carriage and infections keep occurring. Fortunately, outbreaks usually remain limited to a few clustered cases.

**Table 1.**
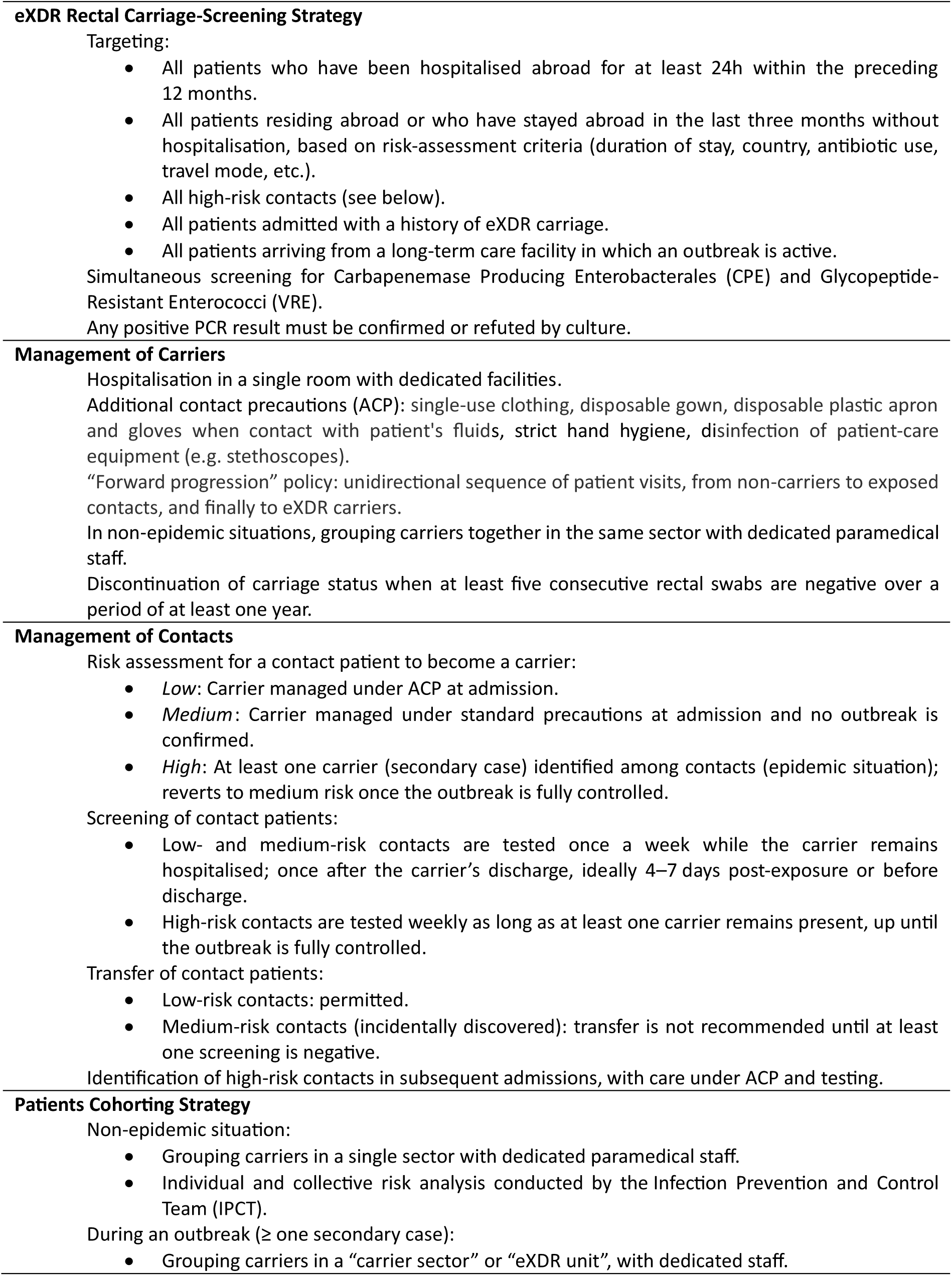
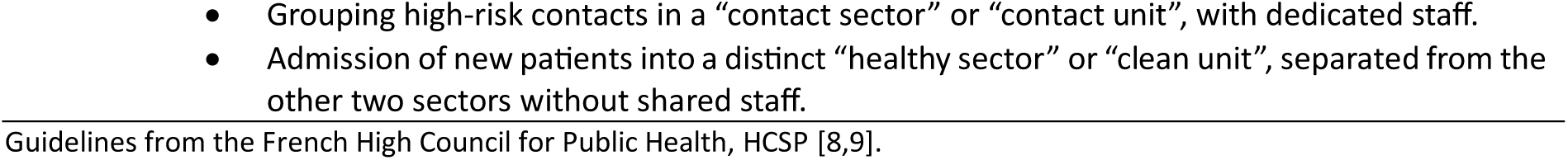
- Main “screen and isolate” control measures of the eXDR carriage outbreak at Hôpital Européen Marseille.

| **eXDR Rectal Carriage-Screening Strategy** |
|---|
| Targeting: |
| • All patients who have been hospitalised abroad for at least 24h within the preceding 12 months. |
| • All patients residing abroad or who have stayed abroad in the last three months without hospitalisation, based on risk-assessment criteria (duration of stay, country, antibiotic use, travel mode, etc.). |
| • All high-risk contacts (see below). |
| • All patients admitted with a history of eXDR carriage. |
| • All patients arriving from a long-term care facility in which an outbreak is active. |
| Simultaneous screening for Carbapenemase Producing Enterobacterales (CPE) and Glycopeptide-Resistant Enterococci (VRE). |
| Any positive PCR result must be confirmed or refuted by culture. |
| **Management of Carriers** |
| Hospitalisation in a single room with dedicated facilities. |
| Additional contact precautions (ACP): single-use clothing, disposable gown, disposable plastic apron and gloves when contact with patient's fluids, strict hand hygiene, disinfection of patient-care equipment (e.g. stethoscopes). |
| “Forward progression” policy: unidirectional sequence of patient visits, from non-carriers to exposed contacts, and finally to eXDR carriers. |
| In non-epidemic situations, grouping carriers together in the same sector with dedicated paramedical staff. |
| Discontinuation of carriage status when at least five consecutive rectal swabs are negative over a period of at least one year. |
| **Management of Contacts** |
| Risk assessment for a contact patient to become a carrier: |
| • *Low*: Carrier managed under ACP at admission. |
| • *Medium*: Carrier managed under standard precautions at admission and no outbreak is confirmed. |
| • *High*: At least one carrier (secondary case) identified among contacts (epidemic situation); reverts to medium risk once the outbreak is fully controlled. |
| Screening of contact patients: |
| • Low- and medium-risk contacts are tested once a week while the carrier remains hospitalised; once after the carrier’s discharge, ideally 4–7 days post-exposure or before discharge. |
| • High-risk contacts are tested weekly as long as at least one carrier remains present, up until the outbreak is fully controlled. |
| Transfer of contact patients: |
| • Low-risk contacts: permitted. |
| • Medium-risk contacts (incidentally discovered): transfer is not recommended until at least one screening is negative. |
| Identification of high-risk contacts in subsequent admissions, with care under ACP and testing. |
| **Patients Cohorting Strategy** |
| Non-epidemic situation: |
| • Grouping carriers in a single sector with dedicated paramedical staff. |
| • Individual and collective risk analysis conducted by the Infection Prevention and Control Team (IPCT). |
| During an outbreak (≥ one secondary case): |
| • Grouping carriers in a “carrier sector” or “eXDR unit”, with dedicated staff. |
| • Grouping high-risk contacts in a “contact sector” or “contact unit”, with dedicated staff. |
| • Admission of new patients into a distinct “healthy sector” or “clean unit”, separated from the other two sectors without shared staff. |

Guidelines from the French High Council for Public Health, HCSP [8,9].

Marseille, France’s second-largest city, is located on the northern shore of the Mediterranean Sea, with a significant proportion of its population regularly travelling to North and sub-Saharan Africa. In early 2025, the Hôpital Européen Marseille (HEM) – a medium-size multidisciplinary hospital – experienced the largest hospital outbreak of eXDR carriage reported to date in France. From January to September 2025, nearly 350 cases of rectal carriage of NDM-producing *E. coli* were identified across all wards. The intensive screening campaign undertaken to contain the outbreak also uncovered numerous sporadic carriers of OXA-48-like-producing Enterobacterales and VRE.

Here, we provide a comprehensive account of this major outbreak. We offer an epidemiological description of its emergence and evolution, and we critically assess the management and encountered challenges. We aim to draw lessons, which may enhance preparedness for managing future eXDR outbreaks.

## 2 Methods

### 2.1 Study Design and Context

We designed a mixed observational study including a retrospective epidemiological analysis of screening data and patients’ movements within HEM from November, 2024 to September, 2025, and a qualitative analysis based on participant observation of the crisis management.

HEM is a roughly 600-bed general hospital located in the centre of Marseille. It is a private non-profit institution participating in the French public hospital healthcare system. The hospital hosts an emergency department and offers all medical specialties except for paediatrics, obstetrics, intracranial neurosurgery, cardiac surgery and solid-organ or bone-marrow transplantation. There is an antimicrobial stewardship program led by infectiologists. Antibiotic use is around national averages in intensive care unit, medicine and surgery wards [13,14].

This non-interventional study was approved by the HEM ethics committee (ref 2025-08-04-SR). The data was processed in compliance with the MR-004 methodology. To preserve confidentiality, dates were replaced by day number, from a Day-0 in late November 2024, when the index case was admitted.

### 2.2 Microbiological analysis

Microbiological analyses were performed by the private laboratory Biogroup (COFRAC accreditation according to the ISO 15189 standard), whose platform is located on site. Rectal swabs for eXDR screening were transported to the laboratory via pneumatic tubes and cultured on chromogenic selective agar plates. These were mostly mSuperCARBA™ medium (CHROMagar, La Plaine Saint-Denis, France) and sometimes CHROMID® Carba Smart medium (bioMérieux, Marcy-l’Étoile, France), depending on availability. Isolates were identified by MALDI-TOF Vitek® MS (bioMérieux) and antimicrobial susceptibility testing was performed on the Vitek® 2 system (bioMérieux). When an enterobacterial isolate was resistant to any carbapenem, a multiplex immunochromatographic NG test® CARBA-5 (NG Biotech, Guipry-Messac, France) was performed; this test was later replaced by PCR. In parallel, multiplex PCR detection of eXDR directly from rectal swabs was used. Nucleic-acid extraction was performed using the Chemagic^TM^ Total NA Extraction Kit H96 (Revvity, Waltham, MA, USA). CPE and VRE detection were performed by multiplex PCR on the CFX Opus system (Bio-Rad France, Marnes-la-Coquette, France) using respectively the CRE ELITe MGB® Kit (ELITechGroup, Torino, Italy) – detecting *bla*_NDM/VIM/IMP_, *bla*_KPC_ and *bla*_OXA-48-like_ genes – and the GeneProof™ VRE PCR Kit (GeneProof, Brno, Czech Republic) – detecting *vanA* and *vanB* genes. From April 1st, 2025, these extraction and PCR kits were replaced by the BD MAX^TM^ system (BD, Franklin Lakes, NJ, USA), detecting *bla*_KPC_, *bla*_NDM_ and *bla*_OXA-48-like_ genes. Throughout the outbreak, a selection of CPE isolates, mostly chosen among new CPE phenotypes or *E. coli* NDM isolates from previously unaffected (non-carrier, non-contact) patients in various wards along the outbreak, was sent to the National Reference Centre for Antibiotic Resistance (NRCAR) for carbapenemase identification and MLST genotyping (Supplementary Table S1).

### 2.3 Qualitative assessment

The HEM infection prevention and control team (IPCT), infectiologists, members of the medical staff committee (CME), as well as microbiologists, senior management, nursing leadership, cohorting-unit staff and housekeeping supervisors contributed to drafting a detailed narrative of the outbreak. Their input enabled the description of the chosen control strategy, including challenges faced at each stage of the outbreak leading to successive adaptations. Data was thematically organised to evaluate the outcome of these control strategies, to identify potential tensions and provide advice to future crisis management summarised in a “lessons learned” box.

### 2.4 Epidemiological Data and Statistical Analysis

Data from eXDR screening tests performed at HEM during the study period was extracted from the laboratory information systems KaliSil® (Dedalus, Florence, Italy) and SIRweb® (i2a, Montpellier, France). The sampling date, techniques used (culture, PCR, or both) and their results were collected, to produce a comprehensive cohort of screened patients. Until the study period, HEM had faced only sporadic cases of eXDR carriage (OXA-48-like and VRE mainly, and isolated cases of NDM-producing *Klebsiella pneumoniae* and *Escherichia coli*), and a small outbreak of OXA-48-like-producing *Enterobacter cloacae* complex in 2024, limited to one unit. During the study period, an NDM-producing *E. coli* carriage was considered as part of the outbreak if it shared the same antimicrobial susceptibility testing profile as the index and previous isolates. In-hospital patients’ movement data during the study period was extracted from the hospital information system Hopital Manager® (Softway Medical, Fuveau, France). The IPCT list of identified and monitored nosocomial contacts was also collected.

#### Initial transmission chains were reconstructed from temporal and spatial data

Nosocomial contacts were retrospectively identified by the computed cross-matching of carriers’ movements with those of other patients in the same care unit (contacts) or in the same room (“super contacts”), irrespective of any acquisition-risk stratification. Epidemic curves describing the evolution of the number of carriers, contacts and infected cases over time were built. Contacts identified by movement analysis were compared with those monitored by the IPCT.

Diagnostic performance of culture versus PCR was evaluated through sensitivity, specificity, positive and negative predictive values, and positive/negative likelihood ratios.

Data was organised in Microsoft Excel 2019®. All statistical analyses were performed in RStudio version 2024.12.1 with R 4.5.0.

## 3 Results

### 3.1 Detection of the Outbreak and Initial Transmission Chain

Early January, 2025, an NDM-producing *E. coli* carrier was identified in a medical ward of HEM. He had been screened 48 hours earlier after contact with a VRE carrier (Figure 1). Three additional NDM-producing *E. coli* carriers hospitalised in 2 other wards were identified 2 days later (patients #3, #4 and #5), also screened after contact with eXDR carriers (Figure 1). The microbiologists immediately alerted the IPCT who declared the outbreak and identified the index carrier (#1), who had been hospitalised in HEM since the end of November, 2024 (Day-0). He had been placed under ACP upon arrival at the emergency department as he had undergone medical care in Senegal before admission, with a rectal screening positive for NDM-producing *E. coli* on the same day (Figure 1). Throughout his stay, the patient remained under ACP; however, due to a shortage of dedicated staff and bed-availability constraints, he could not be assigned to a distinct unit and was transferred 4 times to different units during December 2024, before succumbing to an unrelated cause mid-January 2025. Although non-compliance with the “forward progression” policy (Table 1, [8,9]), his low-risk contacts were screened negative. No clear epidemiological link could be established with patient #2, although patient #1 had been in the same ward 20 days before the arrival of patient #2. Likewise, while a contact transmission might appear plausible between cases #2, #3 and #4, who shared the same unit, patient #5 did not appear linked to them (Figure 1). This raised the possibility of additional unscreened secondary cases, or of an unidentified environmental reservoir.

**Figure 1.**
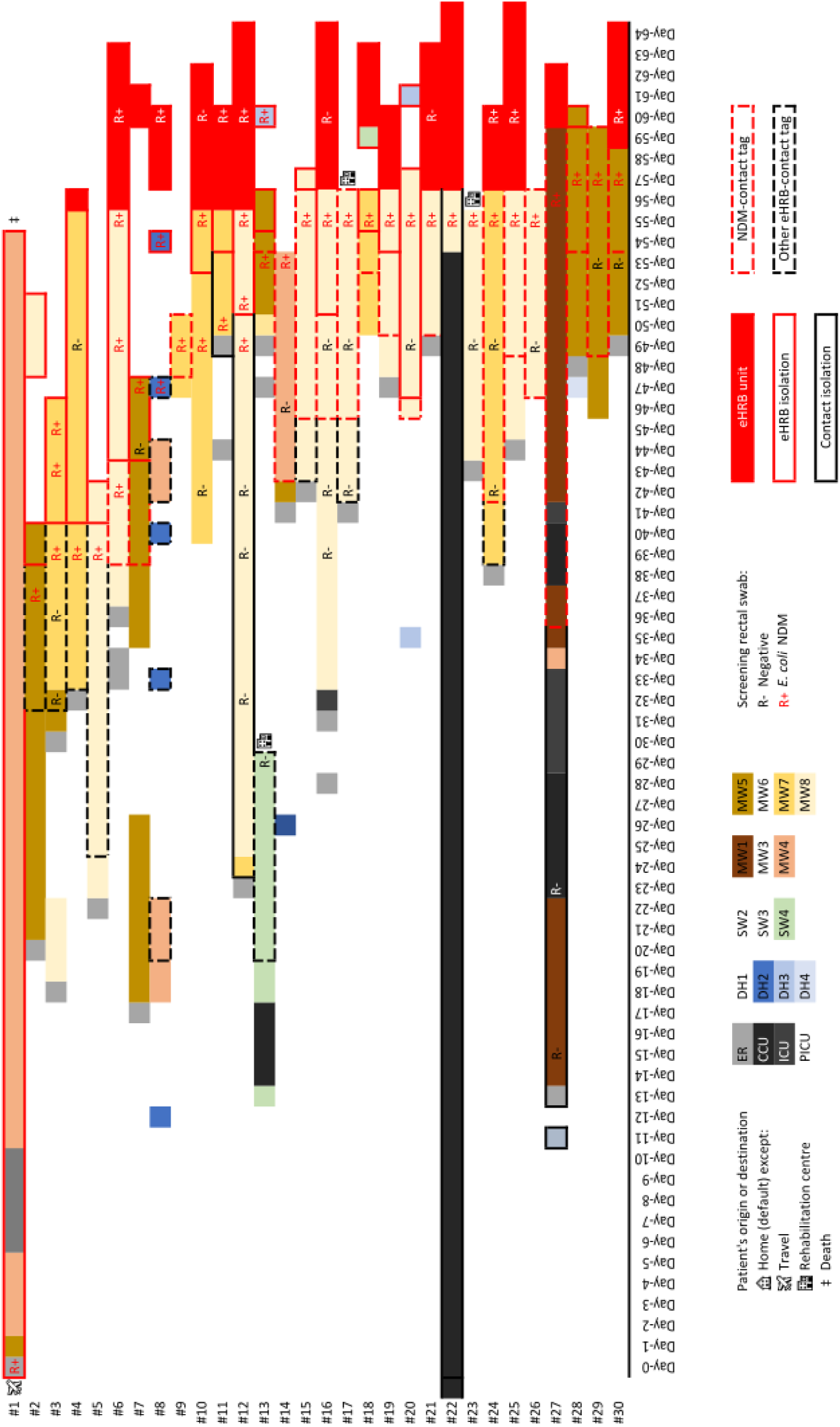
- Gantt chart of the first 30 NDM-producing *Escherichia coli* carriers, Day-0 and Day-64. History of movements within the various inpatient or day-care units in Hôpital Européen Marseille; results of rectal screening; identification as contact; isolation with additional contact precautions; entry and exit routes (excluding patient’s residency); and opportunities for transmission.

Based on the analysis of intra-hospital movements, these five initial NDM-producing *E. coli* carriers could have generated at least 1,178 potential contacts, of which 613 (52 %) were contacts for a period over 24 hours. Among these prolonged contacts, 412 (67 %) occurred before the identification of the outbreak, and 365 (60 %) could be regarded at high-risk of carriage acquisition owing to the absence of ACP applied for patients #2–#5 prior to detection of their carriage.

The spatiotemporal distribution of the first 30 cases delineated three principal clusters in distinct medical wards (MW5, MW7 and MW8). In MW7, the implementation of ACP was delayed (Figure 1). At the time of their positive screening, 11 cases (37 %) had not been recognised as contacts and 21 (70 %) had not been placed under ACP, thereby undoubtedly contributing to the outbreak spread.

### 3.2 Outbreak Management

The IPCT managed the outbreak in accordance with current French national guidelines [8,9] (Table 1), and deployed a substantial proportion of the hospital workforce. A crisis unit, comprising senior hospital managers, infectious disease specialists, chairs of the hospital medical commission and representatives from all clinical departments, laboratory staff, nursing leadership and technical services, convened regularly from Day-41 (Figure 2C). The Regional Health Agency (ARS) was notified in accordance with legal reporting obligations for MDRO to Public Health authorities.

**Figure 2.**
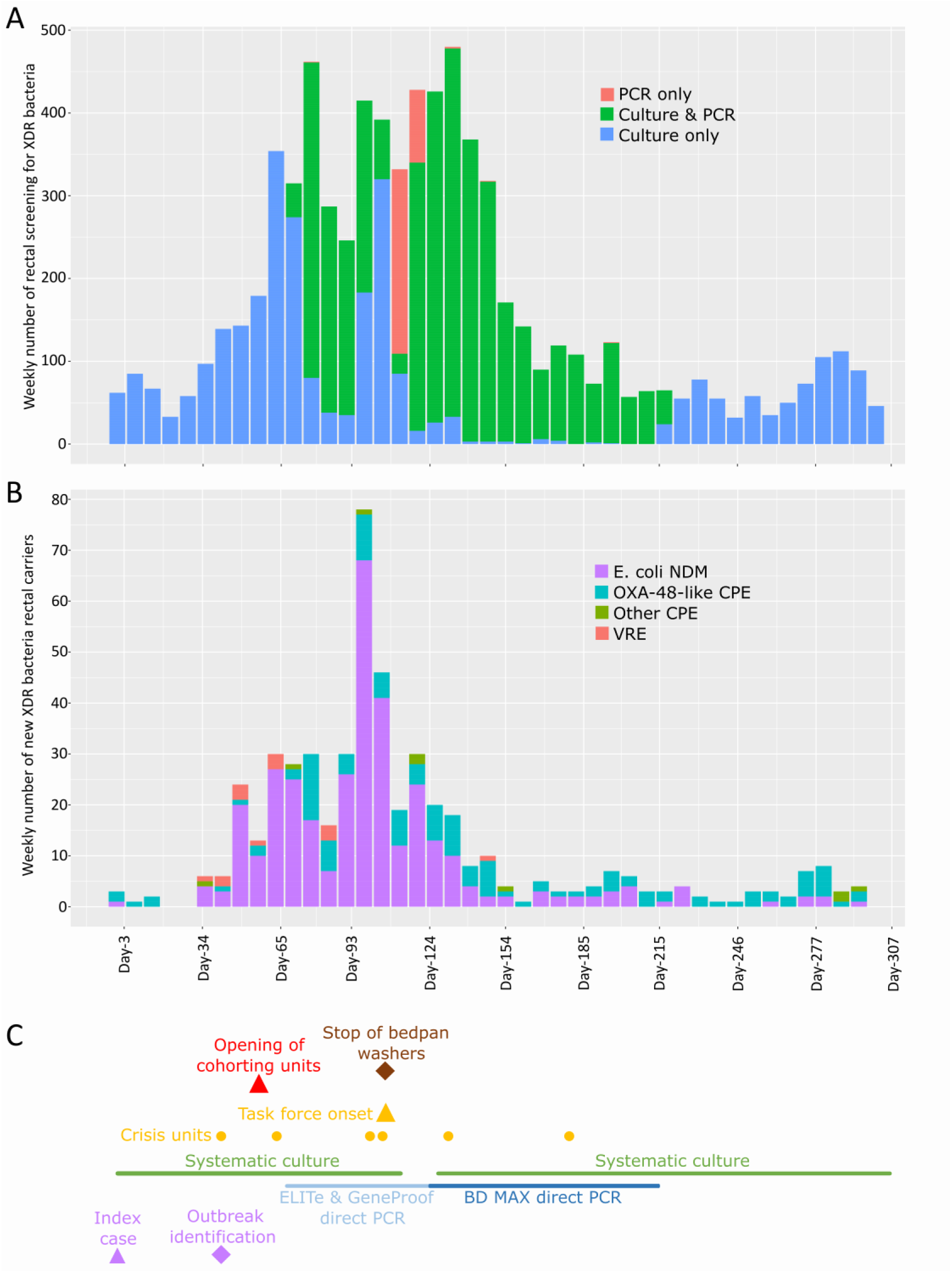
- Weekly trends in screening techniques, bacterial type carriage, and key milestones of the outbreak. (A) Number of rectal tests performed to screen for eXDR carriage, classified by technique; (B) Number of new carriers, classified by bacterial type; and (C) Key milestones of the NDM-producing *Escherichia coli* carriage outbreak. eXDR, extremely drug-resistant bacteria; CPE, carbapenemase-producing Enterobacterales; VRE, vancomycin-resistant *Enterococcus faecium*.

An intensive screening program targeted all contacts – defined as patients admitted in the same care unit as a carrier – using rectal swabbing. In line with recommendations [8,9] (Table 1), contacts were screened on the day of source-patient carriage identification, then on every Tuesday until discharge. Contact patients readmitted to dedicated units were screened on admission and subsequently following the same schedule. Screening of a random sample from apparently unaffected (non-carrier, non-contact) patients was also introduced, which was later expanded to all unaffected patients on Mondays and Thursdays during their first week of admission, then on every Monday. eXDR carriers were screened monthly. Timing adjustments to these procedures were made during the outbreak. At the request of the crisis unit, a direct PCR screening for CPE on rectal swabs without prior culture was introduced on Day-67 (Figure 2A & 2C) to accelerate isolation of new carriers and contacts, at admission and during weekly screening campaigns. The routine culturing of samples was suspended on Day-113 (Figure 2C), but PCR-positive results were still controlled by culture when feasible. Recurrent false negative and positive PCR results using the CRE ELITe MGB® kit, together with difficulties to providing same-day results, prompted a switch to the BD MAX™ system on Day-124, and the reintroduction of the systematic culture of rectal swabs (Figure 2C). Following epidemiological improvement, the high cost of routine screening, the limited operational benefit observed, and suboptimal PCR performance – especially for OXA-48-like isolates (Table 2) – systematic screening of unaffected patients was halted on Day-151. It was replaced by risk-factor-based testing, while contacts were tested only on day 4 and day 7 after exposure.

**Table 2.**
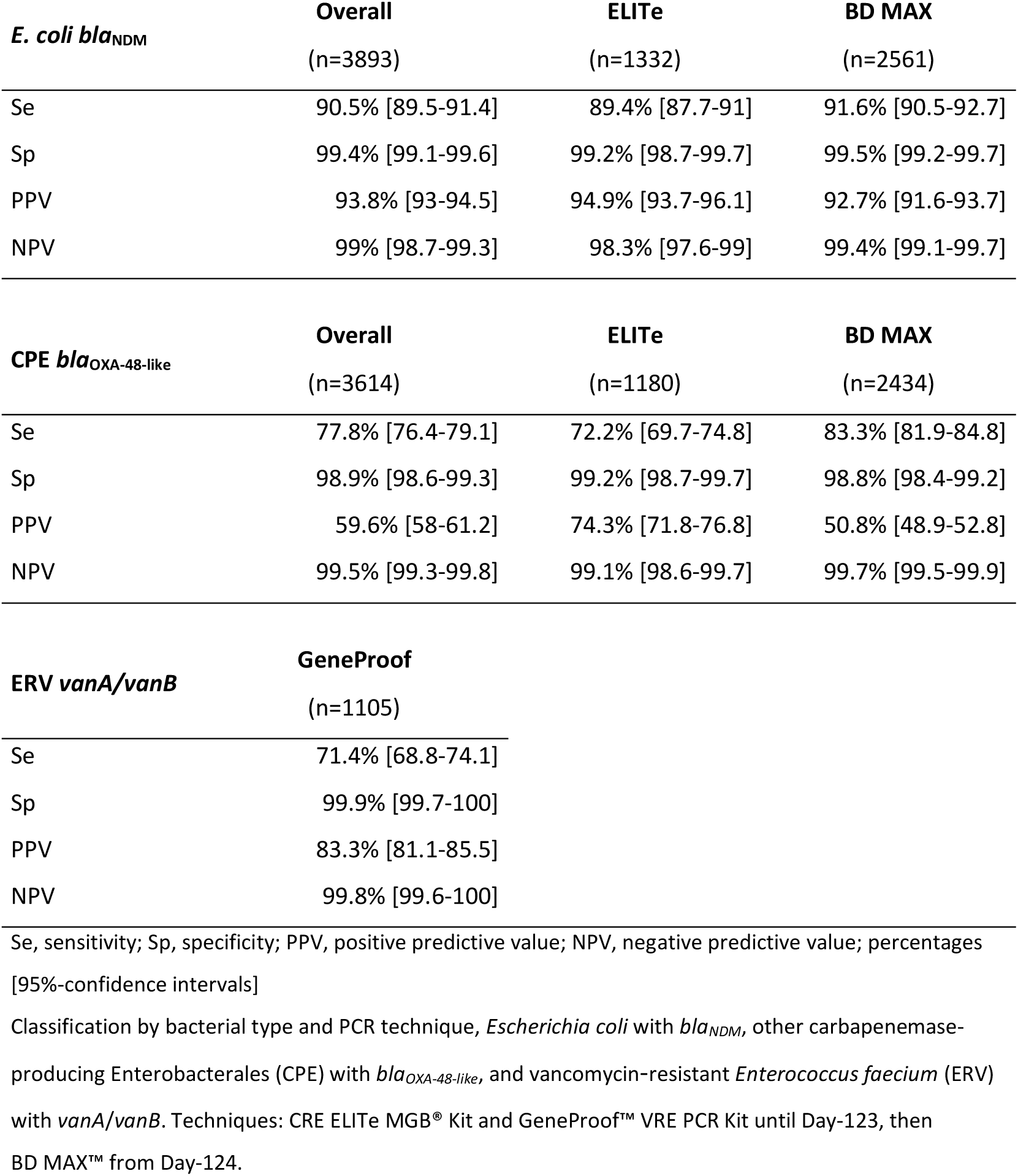
- Performance of direct PCR versus culture for rectal carriage screening of eXDR.

| ***E. coli bla*<sub>NDM</sub>** | **Overall** | **ELITe** | **BD MAX** |
|---|---|---|---|
|  | (n=3893) | (n=1332) | (n=2561) |
| Se | 90.5% [89.5-91.4] | 89.4% [87.7-91] | 91.6% [90.5-92.7] |
| Sp | 99.4% [99.1-99.6] | 99.2% [98.7-99.7] | 99.5% [99.2-99.7] |
| PPV | 93.8% [93-94.5] | 94.9% [93.7-96.1] | 92.7% [91.6-93.7] |
| NPV | 99% [98.7-99.3] | 98.3% [97.6-99] | 99.4% [99.1-99.7] |
| **CPE *bla*<sub>OXA-48-like</sub>** | **Overall** | **ELITe** | **BD MAX** |
|  | (n=3614) | (n=1180) | (n=2434) |
| Se | 77.8% [76.4-79.1] | 72.2% [69.7-74.8] | 83.3% [81.9-84.8] |
| Sp | 98.9% [98.6-99.3] | 99.2% [98.7-99.7] | 98.8% [98.4-99.2] |
| PPV | 59.6% [58-61.2] | 74.3% [71.8-76.8] | 50.8% [48.9-52.8] |
| NPV | 99.5% [99.3-99.8] | 99.1% [98.6-99.7] | 99.7% [99.5-99.9] |
| **ERV *vanA/vanB*** | **GeneProof** |  |  |
|  | (n=1105) |  |  |
| Se | 71.4% [68.8-74.1] |  |  |
| Sp | 99.9% [99.7-100] |  |  |
| PPV | 83.3% [81.1-85.5] |  |  |
| NPV | 99.8% [99.6-100] |  |  |

Se, sensitivity; Sp, specificity; PPV, positive predictive value; NPV, negative predictive value; percentages [95%-confidence intervals]
Classification by bacterial type and PCR technique, *Escherichia coli* with *bla<sub>NDM</sub>*, other carbapenemase-producing Enterobacterales (CPE) with *bla<sub>OXA-48-like</sub>*, and vancomycin-resistant *Enterococcus faecium* (ERV) with *vanA*/*vanB*. Techniques: CRE ELITe MGB® Kit and GeneProof™ VRE PCR Kit until Day-123, then BD MAX™ from Day-124.

From Day-56, patient cohorting was progressively established in three distinct wards (MW3, MW4, then MW1) dedicated to eXDR carriers (Figure 1 & 2C). In accordance with recommendations (Table 1), contacts were also grouped and managed in dedicated sectors to enable the reopening of unaffected areas for new admissions. At the peak of the outbreak, the influx of new contacts and insufficient nursing staff to secure cohorting within units necessitated transferring contacts to dedicated wards.

Throughout the outbreak, extensive on-site day and night investigations were conducted by the IPCT, health administrators and volunteer physicians, and by the Regional Infection Prevention Support Centre (CPias) on three occasions. These investigations consistently revealed breaches in standard hand hygiene practices and ACP (inadequate use of alcohol-based solutions, overuse of single-use gloves, wearing jewellery, no use of disposable gowns and plastic apron, or even visits in street clothes, no disinfection of shared devices, disregard for the “forward progression” policy…). There were also some cohorting errors such as readmission of known contacts to unaffected sectors. These observations suggested primarily contact transmission, prompting significant corrective measures. Mobile teams monitoring units daily listened to the difficulties faced by healthcare staff and reinforced education on best practices. Information sessions were also organised in collaboration with the CPias. Disposable personal protective equipment (single-use gowns, plastic aprons, coveralls) was widely deployed. Emergency and radiology schedules were reorganised to prevent contamination of unaffected patients.

Despite these efforts, new carriers among unaffected patients were identified in decontaminated units, even after displacement of carriers and contacts. This fuelled speculation about the intrinsic transmissibility of this NDM-producing *E. coli* strain or an associated environmental transmission route that could involve any reusable device (such as water dispensers) or a failure in biocleaning (sink and shower siphons disinfection, detergent dilution system). Notably, a major dysfunction in bedpan cleaning was identified. Indeed, in all HEM wards (Supplementary Figure S1), shared polypropylene bedpans were cleaned without prior drainage of excreta using dedicated steam-bath Typhoon® washers (Arjo, Villeneuve d’Ascq, France). All these bedpan washers appeared heavily scale-covered, hindering efficiency, although swabbing to search for eXDR proved negative. Room bio-cleaning was progressively intensified: steam-cleaning; high-level and low-level (floor) surface disinfection; airborne surface disinfection (ASD) using hydrogen peroxide with the Nocolyse® disinfectant and Nocospray® nebulizer (Nosotech, Oxypharm, Champigny-sur-Marne, France) from Day-70; javelisation followed by siphon replacement; and replacement of water dispensers with bottled water. The cessation of reusable bedpans in favour of patient-specific cardboard bedpans covered by a disposable CareBag® bedpan liner with absorbent pad (Cleanis SASU, Arcueil, France) – initially restricted to carrier sectors – was eventually extended hospital-wide from Day-106, pending maintenance and requalification (Figure 2C).

### 3.3 Overview of the Outbreak

Screening rectal swabs were countlessly collected throughout the outbreak: already 984 by the end of January 2025; more than 400 weekly sampled by mid-February and in late March-early April 2025 (Figure 2A); with a cumulative total of 7,486 analysed by September, 2025. This screening strategy led to the identification of an exponentially increasing number of NDM-producing *E. coli* carriers, reaching an initial peak of 27 cases per week in late January 2025. A temporary decline during the second half of February 2025 coincided with a drop in the hospital activity during the winter holidays in France. A second epidemic peak occurred in early March 2025, reaching 68 new cases per week, followed by a gradual decline until the outbreak ended in May 2025. Subsequently, new cases were identified sporadically until the end of September 2025, mostly among contacts (Figure 2B). A total of 343 NDM-producing *E. coli* carriers were recorded, sharing similar antimicrobial susceptibility (only susceptible to aminoglycosides, colistin, fosfomycin and the aztreonam-avibactam combination, and resistant to all other β-lactams including cefiderocol and carbapenems/β-lactamase inhibitors as well as other antibiotic classes including quinolones or sulfamethoxazole). All 14 isolates sent to the NRCAR between January and July 2025 were genotyped MLST ST-44 and carried a *bla*_NDM-5_ gene (Supplementary Table S1).

Out of these 343 identified NDM-producing *E. coli* carriers, 273 (80%) were secondary cases of a likely contact with a previous carrier, retrospectively identified through the computed analysis of hospital movements. The other 20% were considered unaffected (non-carrier, non-contact) prior to detection, including 5% being admitted at HEM for the first time. The entire hospital was concerned by the outbreak, carriers being identified simultaneously in a maximum of 11 wards within the same week at the peak of the outbreak (Figure 3A).

**Figure 3.**
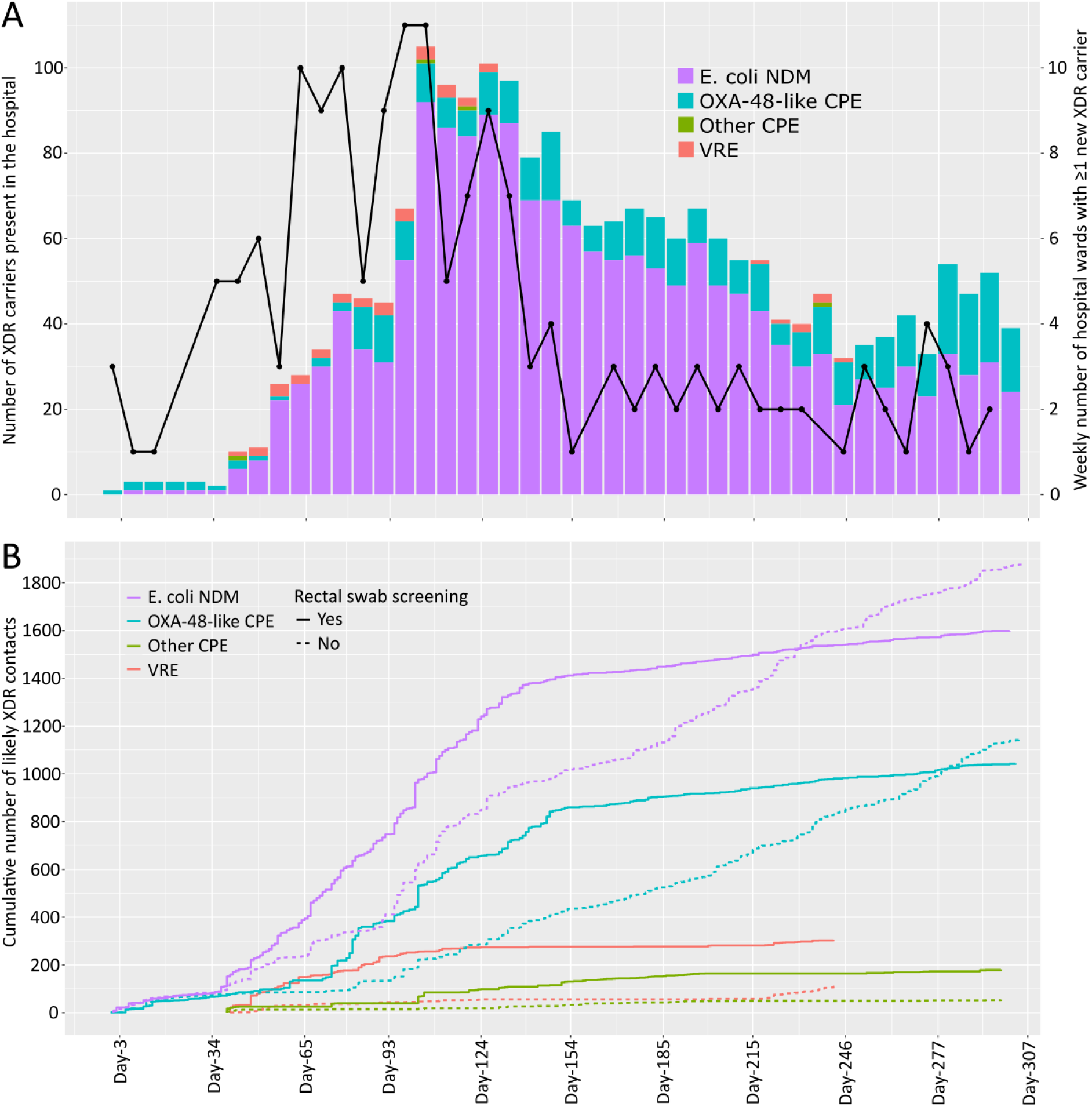
- Evolution of the number of carriers and likely contacts throughout the outbreak. (A) Number of carriers present in various hospital units; (B) Number of units with ≥ 1 new carrier, cumulative number of new contacts lasting ≥24h by bacterial type, and status regarding rectal carriage screening. eXDR, extremely drug-resistant; CPE, carbapenemase-producing Enterobacterales; VRE, vancomycin-resistant *Enterococcus faecium*. Hospital units include inpatient wards, day-care units, emergency department, endoscopy unit, but exclude outpatient consultations.

In addition, a total of 129 OXA-48-like CPE (mostly *K. pneumoniae* strains, mostly carrying *bla*_OXA-48_ and a few *bla*_OXA-244_ or *bla*_OXA-181_, Supplementary Table S1), 9 other CPE isolates (VIM or untyped), and 14 VRE isolates had been identified sporadically by the end of the study period (Figure 2B). Half of these carriages (50%) occurred in *a priori* unaffected patients, indicating their sporadic nature. Among other carriages, several nosocomial secondary transmissions were evidenced by the IPCT and confirmed by genotyping (Supplementary Table S1).

At the height of the outbreak in March 2025, there were more than 90 eXDR carriers (including over 80 NDM carriers) present weekly in HEM (excluding outpatient departments) (Figure 3A).

During the study period, 14 documented infections with NDM-producing *E. coli* were diagnosed in 13 of the 343 carriers (4%), consisting of: 10 urinary tract infections, two biliary-origin bacteraemias, one implant-associated bacteraemia and one primary bacteraemia. Two urinary tract infections occurring in an end-of-life setting were not treated by antimicrobial, and one lower urinary tract infection was treated with nitrofurantoin. The other 11 infections were treated with the antibiotic combination aztreonam-avibactam, five times associating aztreonam with ceftazidime-avibactam (Zavicefta®), and six prescriptions of Emblaveo® through an early access authorisation. These NDM-producing *E. coli* infections were related to deaths of four patients who were otherwise under treatment of life-threatening diseases.

Computed analysis of hospital movements suggested that the NDM-producing *E. coli* carriers had generated 14,262 likely contacts, of whom 3,478 (24%) had been in contact with a carrier for a cumulative duration of at least 24 hours (Figure 3B). Only 2,305 (16%) of them had been tagged as contacts by the IPCT. A total of 2,028 (14%) of computed contacts and 1,608 (70%) of IPCT-tagged contacts were successfully screened, of whom 273 (13%) and 238 (15%) tested positive, respectively. One hundred and five (31%) of all NDM carriers were thus identified by chance through a systematic screening. Of these, 35 (10%) were likely contacts missed by the IPCT tagging and screening.

### 3.4 Real-life PCR performance evaluation

Out of the 7,486 rectal swabs sampled from Day-0 to Day-307, 4,028 (54%) were analysed using both PCR and culture, mainly between February and July 2025 (Figure 2A). NDM-producing *E. coli* was detected by culture in 476 samples, by PCR in 356 (of which 178 by the ELITe technique and 178 by BD MAX), and by both culture and PCR in 332 samples. Taking culture as the reference method, the sensitivity and specificity of PCR for this outbreak NDM-producing *E. coli* strain were 90.5% (i.e., 35 probable false negatives) and 99.4% respectively, and the positive and negative predictive values were 93.8% and 99.0% (Table 2). For OXA-48-like-producing Enterobacterales, the sensitivity and specificity of PCR were 77.8% and 98.9% (38 possible false positives), respectively, and the positive and negative predictive values were 59.6% and 99.5%. For *bla*_NDM,_ the sensitivity of the BD MAX PCR system appeared slightly higher than that of the ELITe kit (91.6% [95% confidence interval: 90.5–92.7] versus 89.4% [87.7–91]). The superiority of the BD MAX system appeared to be greater for *bla*_OXA-48-like_ (sensitivity 83.3% [81.9–84.8] versus 72.2% [69.7–74.8]) (Table 2).

## 4 Discussion

### 4.1 Transmission Routes: Contact and/or Environmental?

This outbreak of gastrointestinal carriage of NDM-producing *E. coli* highlights the CPE propensity to rapidly spread in hospital settings [15,16]. Despite repeated investigations during this outbreak, we could not establish a definitive source or an exclusive mode of transmission – whether contact (obvious gaps in standard precautions and ACP), or environmental. The malfunction of bedpan washers, which was previously reported in the literature as a potential transmission route for VRE [17,18], was strongly suspected of playing a key role in this outbreak. Indeed, until their use was discontinued, 45% of eXDR rectal carriers (data not shown) were classified with a WHO performance status as grade 3 (capable of only limited self-care) or 4 (completely disabled), and incidence swiftly declined afterwards. Yet, effective control of this eXDR carriage outbreak was likely multifactorial, like previously described [19].

The exponential growth of the outbreak also underscores the importance of bacterial inoculum size and the cumulative effect of contacts, even at low risk. This explains why transmission went into overdrive despite implemented measures, which proved unsettling for the hospital staff.

Lessons learned from these investigation limits are summarised in Table 3.

**Table 3.**
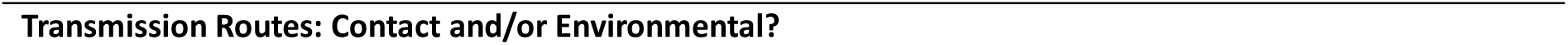
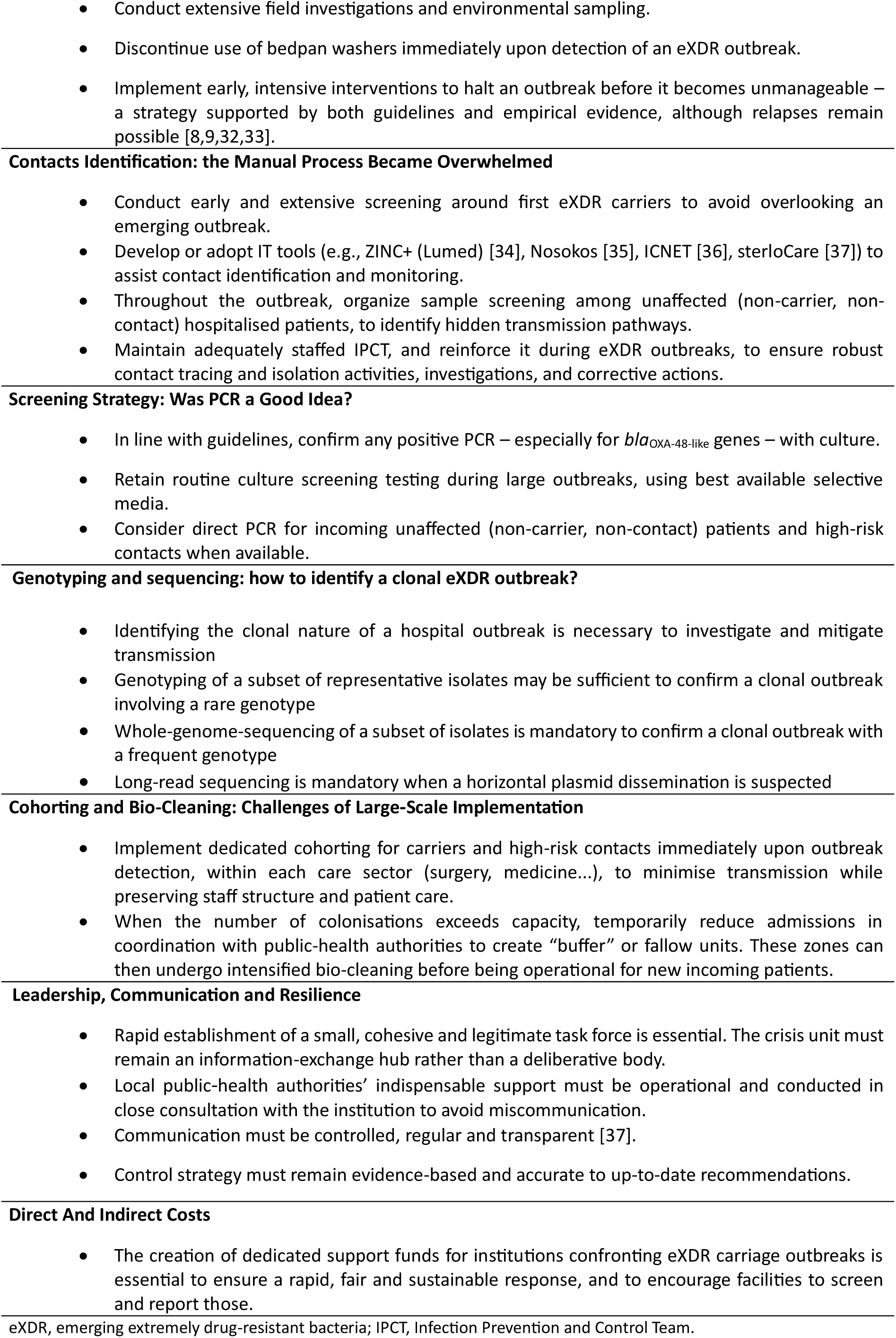
- Lessons learned from this eXDR carriage nosocomial outbreak.

| **Transmission Routes: Contact and/or Environmental?** |
|---|
| • Conduct extensive field investigations and environmental sampling. |
| • Discontinue use of bedpan washers immediately upon detection of an eXDR outbreak. |
| • Implement early, intensive interventions to halt an outbreak before it becomes unmanageable – a strategy supported by both guidelines and empirical evidence, although relapses remain possible [8,9,32,33]. |
| **Contacts Identification: the Manual Process Became Overwhelmed** |
| • Conduct early and extensive screening around first eXDR carriers to avoid overlooking an emerging outbreak. |
| • Develop or adopt IT tools (e.g., ZINC+ (Lumed) [34], Nosokos [35], ICNET [36], sterloCare [37]) to assist contact identification and monitoring. |
| • Throughout the outbreak, organize sample screening among unaffected (non-carrier, non-contact) hospitalised patients, to identify hidden transmission pathways. |
| • Maintain adequately staffed IPCT, and reinforce it during eXDR outbreaks, to ensure robust contact tracing and isolation activities, investigations, and corrective actions. |
| **Screening Strategy: Was PCR a Good Idea?** |
| • In line with guidelines, confirm any positive PCR – especially for *bla*<sub>OXA-48-like</sub> genes – with culture. |
| • Retain routine culture screening testing during large outbreaks, using best available selective media. |
| • Consider direct PCR for incoming unaffected (non-carrier, non-contact) patients and high-risk contacts when available. |
| **Genotyping and sequencing: how to identify a clonal eXDR outbreak?** |
| • Identifying the clonal nature of a hospital outbreak is necessary to investigate and mitigate transmission |
| • Genotyping of a subset of representative isolates may be sufficient to confirm a clonal outbreak involving a rare genotype |
| • Whole-genome-sequencing of a subset of isolates is mandatory to confirm a clonal outbreak with a frequent genotype |
| • Long-read sequencing is mandatory when a horizontal plasmid dissemination is suspected |
| **Cohorting and Bio-Cleaning: Challenges of Large-Scale Implementation** |
| • Implement dedicated cohorting for carriers and high-risk contacts immediately upon outbreak detection, within each care sector (surgery, medicine...), to minimise transmission while preserving staff structure and patient care. |
| • When the number of colonisations exceeds capacity, temporarily reduce admissions in coordination with public-health authorities to create “buffer” or fallow units. These zones can then undergo intensified bio-cleaning before being operational for new incoming patients. |
| **Leadership, Communication and Resilience** |
| • Rapid establishment of a small, cohesive and legitimate task force is essential. The crisis unit must remain an information-exchange hub rather than a deliberative body. |
| • Local public-health authorities’ indispensable support must be operational and conducted in close consultation with the institution to avoid miscommunication. |
| • Communication must be controlled, regular and transparent [37]. |
| • Control strategy must remain evidence-based and accurate to up-to-date recommendations. |
| **Direct And Indirect Costs** |
| • The creation of dedicated support funds for institutions confronting eXDR carriage outbreaks is essential to ensure a rapid, fair and sustainable response, and to encourage facilities to screen and report those. |

eXDR, emerging extremely drug-resistant bacteria; IPCT, Infection Prevention and Control Team.

### 4.2 Contacts Identification: the Manual Process Became Overwhelmed

As the index NDM-producing *E. coli* carrier was immediately isolated under ACP, contacts were deemed low-risk and mostly underwent a single test. This may have hindered secondary cases identification and delayed recognition of the outbreak, which was later incidentally discovered through systematic screening of contacts with other eXDR carriers.

Once the outbreak was declared, the influx of new carriers – including those in previously unaffected wards – made contact identification exceedingly burdensome for the IPCT, even adequately staffed in line with the current French recommendations [20]. Our retrospective computation from patient movement data identified a markedly higher number of contacts than the IPCT, including numerous prolonged contacts (exposure over 24h) and future carriers. These gaps in contact tracing translated into missed opportunities for screening and isolation of potential carriers. They partially explain positive screening in patients classified as unaffected by the IPCT (non-carrier, non-contact), and the overall positivity testing rate (16 %) that far exceeded transmission rates reported in the literature [21]. Of note, the exceptional number of screenings performed during this outbreak identified numerous sporadic non-NDM-producing eXDR carriers, which was unusual in HEM and suggests an underestimated prevalence in the patient population.

Lessons learned from these contact identification challenges are summarised in Table 3.

### 4.3 Screening Strategy: Was PCR a Good Idea?

Since culture-based eXDR detection usually requires up to 3 days, we adopted a mass-screening approach using direct PCR performed on rectal swabs. This was intended to optimise cohorting and rapid decontamination of affected wards, thereby allowing the admission of new unaffected (non-carrier, non-contact) patients. Molecular testing implementation demanded significant organisational effort and equipment from the hospital’s external laboratory partner.

In this real-world setting, the intrinsic performance of direct PCR appeared to be lower than that reported by kit manufacturers, although no discrepancies were identified by the microbiologists in the sample processing and analytical workflow. PCR sensitivity was overall good for blaNDM, and very good with the BD MAX™ system. Sensitivity for blaOXA-48-like genes appeared far below previously published values, especially with the CRE ELITe MGB® Kit [22]. PCR specificity relative to culture appeared very satisfactory, particularly for the outbreak E. coli blaNDM. However, with such a high volume of tests, 38 possible false-positive blaOXA-48-like patients may have been wrongly placed in cohorting units, exposing them to truly acquire an eXDR carriage. Considering that E. coli OXA-244 may be difficult to detect with certain selective culture media such as CHROMID® Carba Smart [23,24], which was marginally used in our laboratory, it is possible that some of the PCR false-positive isolates were in fact PCR true-positives and culture false-negatives. The suboptimal PCR performance, combined with limited operational time savings and the high cost of molecular testing, led us to restrict PCR use to unaffected (non-carrier, non-contact) patients and highest-risk contacts, and to eventually discontinue its use from July 2025. As a matter of fact, a UK modelling study suggested that the cost-benefit balance, in terms of delayed isolation days avoided, also favoured culture over direct PCR [25].

Ultimately, this argues in favour of maintaining routine culture screening using best chromogenic selective agar plates such as mSuperCARBA™ medium (CHROMagar) [24].

Lessons learned from these adjustments in the testing strategy are summarised in Table 3.

### 4.4 Genotyping and sequencing: how to identify a clonal eXDR outbreak?

The occurrence of these clustered cases of rectal carriage of *E. coli* NDM — a CPE strain that had previously only been isolated very rarely within HEM, with no concurrent alerts at other Marseille hospitals — led us to quickly suspect an intrahospital clonal outbreak. All isolates exhibited the same resistance phenotype, most of which were epidemiologically linked, and a probable index case was identified. This reinforced the need to investigate the transmission mechanisms and enabled us to consider possible mitigation strategies.

The results of the NRCAR’s genotyping, showing that 14 of the 15 analysed strains were *E. coli* ST-44 carrying an NDM-5 beta-lactamase, considerably strengthened our suspicion of a clonal outbreak. Indeed, according to NRCAR data, *E. coli* ST-44 was very rare in France in 2024, accounting for 0.64% of the 624 NDM *E. coli* and 0.64% of the 468 NDM-5 *E. coli* genotyped (data not shown, provided by LD from NRCAR). The increase observed in 2025 appears to be largely linked to our outbreak. Under these circumstances, the hypothesis of repeated independent introductions within HEM appears significantly less parsimonious.

Had a much more common *E. coli* genotype been implicated, such as ST-167, whole-genome sequencing (WGS) would have been essential to confirm the clonality of the outbreak. This technique is routinely carried out at the NRCAR when necessary [26].

Finally, none of the isolates’ phenotypes or genotypes suggested horizontal plasmid transmission, the confirmation of which would have required long-read sequencing [26].

Lessons learned about the implications of genotyping and whole-genome sequencing for deciphering a clonal outbreak are summarised in Table 3.

### 4.5 Cohorting and Bio-Cleaning: Challenges of Large Scale Implementation

In line with recommendations [8,9] and based on the risk assessment coordinated by the IPCT, the index case – travelling from Senegal where NDM-producing *E. coli* is emerging [27] – was isolated under ACP in a single-occupancy room but was not placed in a unit with dedicated staff. Once the outbreak was declared, cohorting of eXDR carriers and high-risk contacts was gradually introduced. However, the eXDR cohorting unit could only become operational two weeks later. At the peak of the outbreak, implementation of this cohorting encountered numerous obstacles, some of which have already been documented in the literature [28]: inability to sort carriers by eXDR type – leading to high risk of cross-transmission within eXDR units; repeated in-hospital transfers of initially unaffected patients, who became contacts then carriers; hasty bio-cleaning of rooms and wards – aiming to free up wards for new admissions via the emergency room or scheduled pathways in a high-throughput hospital, leading to cleaning staff exhaustion and cross-contaminations; disruption of healthcare teams – in requisitioned units for cohorting, disturbing routine medical care and therefore the quality of patient management [29]; or strategic inconsistencies in bio-cleaning protocols.

Lessons learned from these limits in the cohorting and bio-cleaning recommendations are summarised in Table 3.

### 4.6 Leadership, Communication and Resilience

This eXDR-carriage hospital outbreak reminded the staff of the past COVID-19 crisis in terms of uncertainty and lack of preparedness. Nevertheless, the regional context was profoundly different, characterised by an institutional isolation, given that the outbreak was confined to HEM. Unlike the pandemic, there were only a few infections related to the outbreak, and the focus was rather given on asymptomatic carriage. There were some limitations in the reaction of regional public-health authorities: a delayed recognition of the outbreak, a lack of operational support. The hospital also experienced ostracism from other facilities, notably medical and rehabilitation centres, which limited the admission of HEM-originating patients.

Internally, cohesion was compromised: although management teams and technical services displayed remarkable mobilisation, the implementation of a structured medical strategy was delayed. Indeed, the crisis unit turned into an ineffective deliberation forum without decision-making conclusions, and poorly coordinated or even contradictory announcements were made by the different stakeholders – senior management, the IPCT, or the Regional Infection Prevention Support Centre (CPias). Senior management had sometimes to make operational decisions urgently, without prior collegial validation. Beliefs and proposals not grounded in evidenced-based medicine were spread by some healthcare workers. Notably, the suspicion of a wider undetected eXDR circulation among other institutions in Marseille – which was later disproved – led some physicians to advocate the cessation of screening; carrier decontamination with colistin and gentamicin oral intake was requested by other clinicians, although not recommended [30]. These difficulties likely generated escalating tensions and, at times, eroded confidence in the control strategy. They strained the crisis-management actors in the context of staff shortages and an extensive crisis: the IPCT (who struggled to enforce directives and faced indifference or even denial, contributing to the resignation of the IPCT pharmacist); senior management (who lacked a director during the outbreak); the bio-cleaning staff, nurses facing fragmented care; and physicians compelled to visit patients scattered across multiple units.

A turning point occurred mid-March 2025 after the formation of a small task force of fewer than 10 persons, including the infection-control pharmacist, infectiologists, senior management and chairs of the medical staff committee. This tighter group clarified decisions and restored coherence in communication. The crisis unit became a structured information forum, no longer a decision-making venue. A legacy of the COVID-19 crisis – a large WhatsApp group comprising over 170 hospital physicians – was reactivated to exchange essential information, including donning procedures, evolving care-unit mapping and antibiotic or proton-pump inhibitor stewardship recommendations. A bed-blocker cell, comprising responsible physicians, leadership, social services and the IPCT, was established to facilitate transfer or discharge of carriers.

Lessons learned from these leadership and communication issues are summarised in Table 3.

### 4.7 Direct and Indirect Costs

The total financial impact of this outbreak on HEM’s resources was estimated at €1.8 million, roughly €5 000 per NDM-producing *E. coli* carrier – a figure comparable to that reported by three Parisian hospitals in 2016 [31]. Direct cost overruns associated with staff, consumables, equipment and bio-cleaning procedures (notably 1,285 ASD by end of September 2025) were compounded by substantial intangible losses: loss of revenue from bed-blockers or unusable double rooms, reputational damage, and admission cancellations by patients or clinicians.

Although the epidemiological risk far exceeded HEM boundaries, stark limits emerged in the inter-hospital solidarity and in the authority of the Regional Health Agency (ARS) regarding the regional structuring of care for eXDR carriers, particularly concerning transfer to rehabilitation centres of patients who had become bed-blockers. In March 2026, the ARS eventually granted €500 000 financial compensation to HEM – a private institution subject to financial balance obligations yet participating in the Public Hospital service. Indeed, loss of operational capacity was an issue for the continuity of care. A claim for loss of revenue is still under assessment by the hospital’s insurer and has raised complex legal matters concerning the definition of an outbreak (asymptomatic carriage is not a disease; can an outbreak be limited to one hospital?).

Lessons learned from the financial impact of this outbreak are summarised in Table 3.

## 5 Conclusion

Despite the unstructured collection of qualitative data, this mixed-method study provides a first-hand account of managing an in-hospital outbreak of eXDR carriage. It demonstrates that, even when transmission mechanisms are difficult to confirm, it is essential to counter contact transmission while simultaneously securing the handling of excreta through the systematic utilisation of single-use bedpans. Rapid PCR screening performed directly on rectal swabs can be operationally useful, provided results are available within a few hours. However, the test is costly, and its imperfect performance may lead to erroneous cohorting of patients. In a large-scale outbreak, cohorting may lead to a substantial reduction in activity and bed closures, necessitating coordination between public-health authorities and clinical teams. The resulting loss of activity and significant costs overruns warrant the use of appropriate indemnity mechanisms. Despite its slow progression, such a carriage crisis requires legitimate and tightly focused governance, together with clear and targeted communication – essential conditions to ensure the effectiveness of interventions and to preserve team resilience.

Despite numerous obstacles, this study illustrates how, five years after the COVID-19 crisis, a first-line hospital was able to halt, within six months, the largest eXDR carriage nosocomial outbreak ever described in France, with the hope of contributing to a better preparedness in the future.

## Data Availability

All data produced in the present study are available upon reasonable request to the authors.

## Authors’ contributions

Study design: SR. Data collection: SR, FSS, JB. Data analysis: SR, VP. Results interpretation: SR, ALH, FSS, CG, NB, CS, LM, LC, CF, MC, JB. Manuscript drafting: SR. Manuscript reviewing: ALH, VP, FSS, CG, NB, CS, LM, LC, CF, LD, MC, JB.

## Acknowledgments

Authors are deeply thankful to the bio-cleaning, nursing, medical, laboratory, technical and management staff involved in this outbreak. They thank Yassine Alibouch, microbiologist at the laboratory, who provided data.

## Supplementary figure and table

**Supplementary Figure S1.**
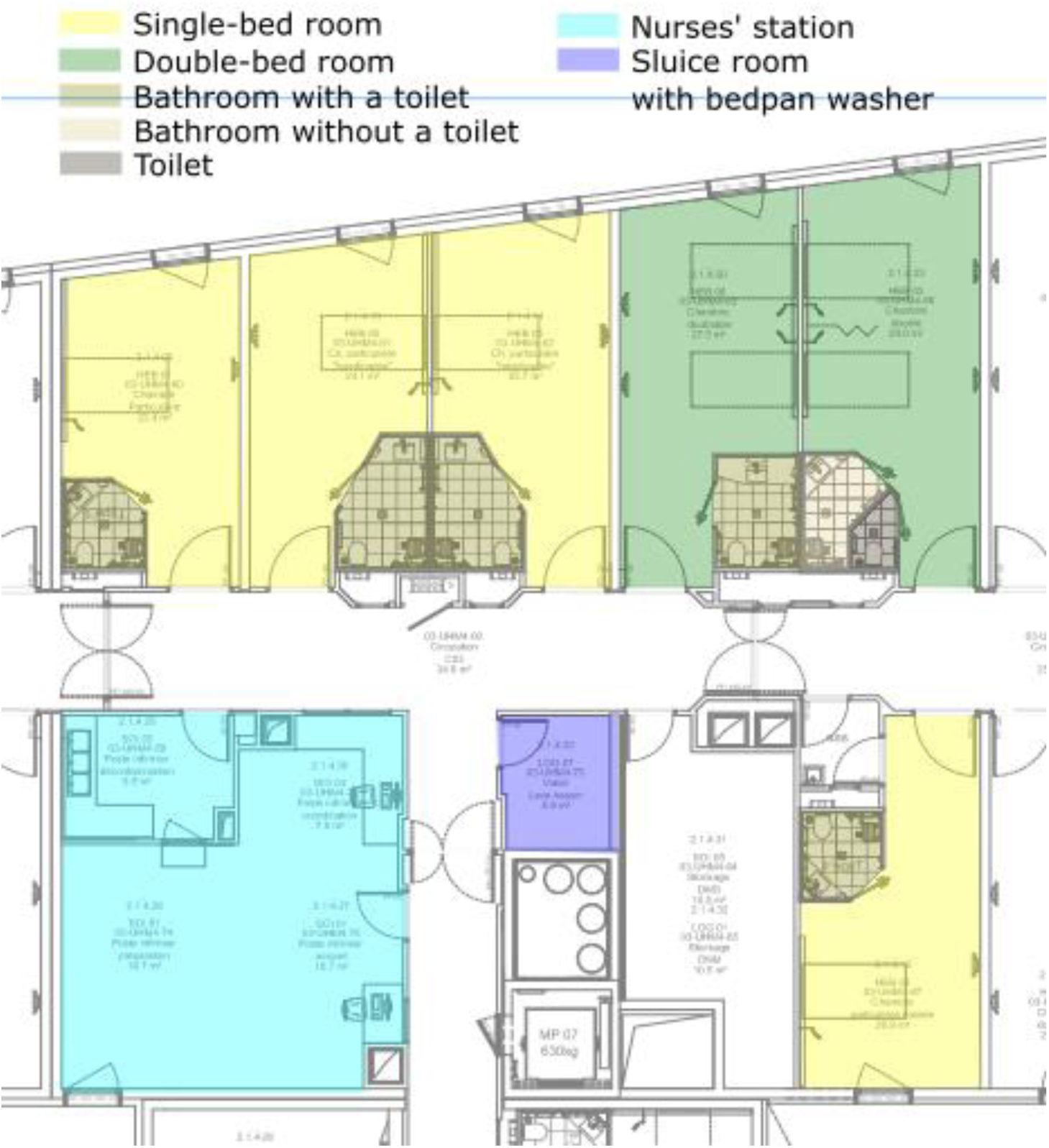
- Floor plan of a section of a typical hospital ward at Hôpital Européen Marseille. The map shows the layout of single and double rooms, bathrooms and toilets, the nurses’ station, and the sluice room where the bedpan washer is installed.

**Supplementary Table S1.** – List of carbapanem-resistant *Enterobacterales* isolates sent to the French National Reference Centre for Antibiotic Resistance (NRCAR) for typing in 2024-2025, including the antimicrobial susceptibility testing result at Biogroup, betalactamase type and MLST genotyping. Colors show isolates of the *Escherichia coli* NDM outbreak in purple, isolates of a limited *K. pneumoniae* OXA-48 outbreak in blue, and isolates of a limited *E. coli* OXA-244 outbreak in green. Sporadic eXDR isolates are not coloured.

| Sampling day* | Patient ID | Sample type | Species | eXDR type | MLST genotype | Ampicillin | Amoxicillin-clavulanate | Cefotaxim | Ceftazidim | Cefoxitin | Ticarcillin | Piperacillin-tazobactam | Ertapenem | Imipenem | Amikacin | Gentamicin | Tobramycin | Ciprofloxacin | Levofloxacin | Cotrimoxazole | AST profile number |
|---|---|---|---|---|---|---|---|---|---|---|---|---|---|---|---|---|---|---|---|---|---|
| -178 | 2 | Backbone | E. cloacae complex | OXA-48 | ST-1483 | R | R | R | R | R | R | R | R | R | R | R | R | R |  | R | 24 |
| -174 | 3 | Rectal swab | E. coli | NDM-5 | ST-1485 | R | R | R | R | R | R | R | R | R | S | S | S | S |  | R | 39 |
| -149 | 4 | Rectal swab | K. pneumoniae | NDM-7 | ST-1296 | R | R | R | R |  | R | R | R | R | S | R | R | S |  | R | 39 |
| -42 | 5 | Rectal swab | E. coli | NDM-5 | ST-4178 | R | R | R | R | R | R | R | R | R | S | S | S | S | S | S | 40 |
| 12 | 6 | Rectal swab | E. coli | OXA-181 | ST-540 | R | R | R | R |  | R | R | R | S | S | S | S | R | R | S | 38 |
| 37 | 7 | Rectal swab | E. coli | NDM-5 | ST-44 | R | R | R | R | R | R | R | R | R | S | S | S | R | R | R | 37 |
| 39 | 8 | Rectal swab | E. coli | NDM-5 | ST-44 | R | R | R | R | R | R | R | R | R | S | S | S | R | R | R | 37 |
| 39 | 9 | Rectal swab | E. coli | NDM-5 | ST-44 | R | R | R | R | R | R | R | R | R | S | S | S | R | R | R | 37 |
| 39 | 10 | Rectal swab | E. coli | NDM-5 | ST-44 | R | R | R | R | R | R | R | R | R | S | S | S | R | R | R | 37 |
| 42 | 11 | Rectal swab | E. coli | NDM-5 | ST-44 | R | R | R | R | R | R | R | R | R | S | S | S | R | R | R | 37 |
| 48 | 12 | Rectal swab | C. freundii | VIM-1 + VIM-4 | ST-98 | R | R | R | R | R | R | R | R | R | S | S | R | R | R | S | 38 |
| 49 | 13 | Rectal swab | E. coli | NDM-5 | ST-44 | R | R | R | R | R | R | R | R | R | S | S | S | R | R | R | 37 |
| 49 | 14 | Rectal swab | E. coli | NDM-5 | ST-44 | R | R | R | R | R | R | R | R | R | S | S | S | R | R | R | 37 |
| 49 | 15 | Rectal swab | E. coli | NDM-5 | ST-44 | R | R | R | R | R | R | R | R | R | S | S | S | R | R | R | 37 |
| 50 | 16 | Rectal swab | K. pneumoniae | OXA-48 | ST-834 | R | R | R | R | S | R | R | R | S | S | S | S | R | R | S | 38 |
| 52 | 17 | Rectal swab | E. coli | NDM-5 | ST-44 | R | R | R | R | R | R | R | R | R | S | S | S | R | R | R | 37 |
| 60 | 18 | Rectal swab | E. coli | OXA-181 | ST-8489 | R | R | R | S | S | R | R | R | S | S | S | S | R | R | S | 57 |
| 60 | 19 | Rectal swab | E. coli | OXA-48 | ST-1598 | R | R | R | R | S | R | R | R | S | S | S | S | I | S | R | 39 |
| 81 | 20 | Rectal swab | K. pneumoniae | OXA-48 | ST-632 | R | R | R | R | S | R | R | R | S | S | S | S | R | R | R | 37 |
| 81 | 21 | Rectal swab | K. pneumoniae | OXA-48 | ST-632 | R | R | R | R | R | R | R | R | S | S | R | R | R | R | R | 37 |
| 81 | 22 | Rectal swab | K. pneumoniae | OXA-48 | ST-632 | R | R | R | R | S | R | R | R | S | S | S | S | R | R | R | 37 |
| 81 | 23 | Rectal swab | K. pneumoniae | OXA-48 | ST-632 | R | R | R | R | S | R | R | R | R | S | S | S | R | R | R | 37 |
| 81 | 24 | Rectal swab | K. pneumoniae | OXA-48 | ST-632 | R | R | R | R | S | R | R | R | S | S | S | S | R | R | R | 37 |
| 88 | 25 | Rectal swab | K. pneumoniae | OXA-48 | ST-632 | R | R | R | R | S | R | R | R | R | S | S | S | R | R | R | 37 |
| 103 | 26 | Rectal swab | E. coli | NDM-5 | ST-6260 | R | R | R | R | R | R | R | R | S |  | R | R | R | R | R | 14 |
| 103 | 27 | Rectal swab | E. coli | OXA-181 | ST-648 | R | R | R | R | R | R | R | R | I |  | R | R | R | R | R | 14 |
| 103 | 28 | Rectal swab | E. coli | OXA-244 | ST-394 | R | R | R | S | S | R | R | R | I | S | S | S | S | S | S | 59 |
| 103 | 29 | Rectal swab | E. coli | OXA-244 | ST-394 | R | R | R | S | S | R | R | R | I | S | S | S | S | S | S | 59 |
| 103 | 30 | Rectal swab | E. coli | OXA-244 | ST-394 | R | R | R | S | S | R | R | R | I | S | S | S | S | S | S | 59 |
| 103 | 31 | Rectal swab | E. coli | OXA-244 | ST-394 | R | R | R | S | S | R | R | R | I | S | S | S | S | S | S | 59 |
| 109 | 32 | Rectal swab | E. coli | NDM-5 | ST-44 | R | R | R | R | R | R | R | R | R | S | S | S | R | R | R | 37 |
| 109 | 33 | Rectal swab | E. coli | NDM-5 | ST-44 | R | R | R | R | R | R | R | R | R | S | S | S | R | R | R | 37 |
| 109 | 25 | Rectal swab | E. coli | NDM-5 + OXA-48 | ST-44 | R | R | R | R | R | R | R | R | R | S | S | R | R | R | R | 37 |
| 131 | 34 | Rectal swab | E. coli | NDM-5 | ST-44 | R | R | R | R | R | R | R | R | R | S | S | S | R | R | R | 37 |
| 221 | 35 | Rectal swab | E. coli | NDM-5 | ST-44 | R | R |  | R | R | R | R | R | R | S | S | S |  | R | R | 37 |
| 280 | 36 | Rectal swab | K. pneumoniae | NDM-5 | ST-147 | R | R |  | R | R | R | R | R | R | R | S | R |  | R | R | 24 |

\* sampling day relative to outbreak onset
E. coli, Escherichia coli; E. cloacae, Enterobacter cloacae; K. pneumoniae, Klebsiella pneumoniae; C. freundii, Citrobacter freundii
S, susceptible ; I, susceptible, increased exposure ; R, resistant

## References

1. World Health Organization. Global antibiotic resistance surveillance report 2025: WHO Global Antimicrobial Resistance and Use Surveillance System (GLASS). Geneva: WHO; 2025 p. 114. https://www.who.int/publications/i/item/9789240116337. Accessed 3 Mar 2026

2. Naghavi M, Vollset SE, Ikuta KS, Swetschinski LR, Gray AP, Wool EE, et al. Global burden of bacterial antimicrobial resistance 1990–2021: a systematic analysis with forecasts to 2050. The Lancet; 2024;0. 10.1016/S0140-6736(24)01867-1

3. World Health Organization. WHO Bacterial Priority Pathogens List, 2024: bacterial pathogens of public health importance to guide research, development and strategies to prevent and control antimicrobial resistance. Geneva: WHO; 2024 p. 72. https://www.who.int/publications/i/item/9789240093461. Accessed 3 Mar 2026

4. Macesic N, Uhlemann A-C, Peleg AY. Multidrug-resistant Gram-negative bacterial infections. The Lancet; 2025;405:257–72. 10.1016/S0140-6736(24)02081-6

5. European Centre for Disease Prevention and Control (ECDC). Surveillance Atlas of Infectious Diseases. https://atlas.ecdc.europa.eu/public/index.aspx. Accessed 14 Apr 2026

6. Tryfinopoulou K, Linkevicius M, Pappa O, Alm E, Karadimas K, Svartström O, et al. Emergence and persistent spread of carbapenemase-producing Klebsiella pneumoniae high-risk clones in Greek hospitals, 2013 to 2022. Eurosurveillance. European Centre for Disease Prevention and Control; 2023;28:2300571. 10.2807/1560-7917.ES.2023.28.47.2300571

7. European Centre for Disease Prevention and Control. Increase in Escherichia coli isolates carrying blaNDM-5 in the European Union/European Economic Area, 2012–2022. Stockholm: ECDC; 2023 May p. 23. 10.2900/72700

8. Lepelletier D, Berthelot P, Lucet J-C, Fournier S, Jarlier V, Grandbastien B. French recommendations for the prevention of ‘emerging extensively drug-resistant bacteria’ (eXDR) cross-transmission. Journal of Hospital Infection; 2015;90:186–95. 10.1016/j.jhin.2015.04.002

9. Haut Conseil de la Santé Publique. Actualisation des recommandations relatives aux BHRe [Internet]. Rapport de l’HCSP. Paris: HCSP; 2019 Dec p. 101. https://www.hcsp.fr/explore.cgi/avisrapportsdomaine?clefr=758. Accessed 3 Mar 2026

10. World Health Organization. Transmission-based precautions for the prevention and control of infections: aide-memoire. WHO; 2022 June p. 4. https://www.who.int/publications/i/item/WHO-UHL-IHS-IPC-20222. Accessed 24 Mar 2026

11. Coppéré Z, Voiriot G, Blayau C, Gibelin A, Labbe V, Fulgencio JP, et al. Disparity of the “screen-and-isolate” policy for multidrug-resistant organisms: A national survey in French adult ICUs. Am J Infect Control. 2018;46:1322–8. 10.1016/j.ajic.2018.05.025

12. Colomb-Cotinat M, Soing-Altrach S, Leon A, Savitch Y, Poujol I, Naas T, et al. Emerging extensively drug-resistant bacteria (eXDR) in France in 2018. Médecine et Maladies Infectieuses. 2020;50:715–22. 10.1016/j.medmal.2020.01.011

13. La surveillance en réseau de la consommation d’antibiotiques et des résistances bactériennes. ConsoRes. https://www.consores.fr/. Accessed 14 Apr 2026

14. SPF. Surveillance de la consommation d’antibiotiques et des résistances bactériennes en établissement de santé. Mission SPARES. Principaux résultats 2024 [Internet]. Saint-Maurice: Santé publique France; 2025 Nov p. 12. https://www.santepubliquefrance.fr/import/surveillance-de-la-consommation-d-antibiotiques-et-des-resistances-bacteriennes-en-etablissement-de-sante.-mission-spares.-principaux-resultats-2024. Accessed 3 Mar 2026

15. Elton L, Williams A, Ali S, Heaphy J, Pang V, Commins L, et al. Tracing the transmission of carbapenem-resistant Enterobacterales at the patient: ward environmental nexus. Ann Clin Microbiol Antimicrob. 2024;23:108. 10.1186/s12941-024-00762-8

16. Robert M, Corvec S, Andreo A, Gallou FL, Marquot G, Mangeant R, et al. Epidemiological and bacteriological trends from 2013 to 2023 of carbapenemase-producing enterobacterales (CPE) in a French university hospital: A permanent risk of outbreak. Infect Dis Now. 2025;55:105021. 10.1016/j.idnow.2024.105021

17. Bryce E, Lamsdale A, Forrester L, Dempster L, Scharf S, McAuley M, et al. Bedpan washer disinfectors: an in-use evaluation of cleaning and disinfection. Am J Infect Control. 2011;39:566–70. 10.1016/j.ajic.2010.10.028

18. Chadwick P, Oppenheim B. Vancomycin-resistant enterococci and bedpan washer machines. The Lancet; 1994;344:685. 10.1016/S0140-6736(94)92119-9

19. French CE, Coope C, Conway L, Higgins JPT, McCulloch J, Okoli G, et al. Control of carbapenemase-producing Enterobacteriaceae outbreaks in acute settings: an evidence review. J Hosp Infect. 2017;95:3–45. 10.1016/j.jhin.2016.10.006

20. Société Française d’Hygiène Hospitalière. Les éléments clés des programmes de prévention et controle des infections (PCI) dans les établissements de santé et médico-sociaux. Rôle, missions, moyens et effectifs des équipes opérationnelles d’hygiène et des équipes mobiles d’hygiène. SF2H; 2022 Feb p. 25. https://www.sf2h.net/actualites/rapport-sf2h-role-missions-moyens-et-effectifs-des-eoh-et-emh.html. Accessed 28 Apr 2026

21. Chang E, Chang HE, Shin IS, Oh YR, Kang CK, Choe PG, et al. Investigation on the transmission rate of carbapenemase-producing carbapenem-resistant Enterobacterales among exposed persons in a tertiary hospital using whole-genome sequencing. J Hosp Infect. 2022;124:1–8. 10.1016/j.jhin.2022.03.005

22. Noster J, Thelen P, Hamprecht A. Detection of Multidrug-Resistant Enterobacterales—From ESBLs to Carbapenemases. Antibiotics. Multidisciplinary Digital Publishing Institute; 2021;10:1140. 10.3390/antibiotics10091140

23. Hoyos-Mallecot Y, Naas T, Bonnin RA, Patino R, Glaser P, Fortineau N, et al. OXA-244-Producing Escherichia coli Isolates, a Challenge for Clinical Microbiology Laboratories. Antimicrobial Agents and Chemotherapy. American Society for Microbiology; 2017;61:10.1128/aac.00818-17. https://doi.org/10.1128/aac.00818-17

24. Emeraud C, Biez L, Girlich D, Jousset AB, Naas T, Bonnin RA, Dortet L. Screening of OXA-244 producers, a difficult-to-detect and emerging OXA-48 variant? J Antimicrob Chemother 2020 Aug 1;75(8):2120–2123. doi: 10.1093/jac/dkaa155.

25. Knight GM, Dyakova E, Mookerjee S, Davies F, Brannigan ET, Otter JA, et al. Fast and expensive (PCR) or cheap and slow (culture)? A mathematical modelling study to explore screening for carbapenem resistance in UK hospitals. BMC Med. 2018;16:141. 10.1186/s12916-018-1117-4

26. Ménard G, Huart D, Le Carrour-Quéré K, Gourmelen F, Le Bihan AM, Guérin F, et al. Long-term dissemination of NDM-producing Enterobacterales in two French hospitals (2020–2024): outbreak complexity and the role of drains as environmental reservoirs. Antimicrob Resist Infect Control. 2026;15:100. 10.1186/s13756-026-01766-z

27. Sarr H, Niang AA, Diop A, Mediannikov O, Zerrouki H, Diene SM, et al. The Emergence of Carbapenem- and Colistin-Resistant Enterobacteria in Senegal. Pathogens. Basel, Switzerland; 2023;12:974. 10.3390/pathogens12080974

28. Legeay C, Thépot-Seegers V, Pailhoriès H, Hilliquin D, Zahar J-R. Is cohorting the only solution to control carbapenemase-producing Enterobacteriaceae outbreaks? A single-centre experience. J Hosp Infect. 2018;99:390–5. 10.1016/j.jhin.2018.02.003

29. Matt M, Senard O, Deconinck L, Lawrence C, Dinh A, Godin E, et al. État des lieux de la perte de chance liée au *cohorting* des patients colonisés et/ou infectés à BHRe en secteur dédié de maladies infectieuses. Médecine et Maladies Infectieuses. 2017;47:S32. 10.1016/j.medmal.2017.03.077

30. Rodríguez Feria D, Diaz Brochero CR, Muñoz Velandia O, Verhelst López JM, Garzón Herazo JR. Effectiveness and safety of oral antibiotics as a decolonization strategy for carbapenem-resistant Enterobacteriaceae: A systematic review of randomized and non-randomized studies. Infectious Diseases Now. 2025;55:105080. 10.1016/j.idnow.2025.105080

31. Birgand G, Leroy C, Nerome S, Luong Nguyen LB, Lolom I, Armand-Lefevre L, et al. Costs associated with implementation of a strict policy for controlling spread of highly resistant microorganisms in France. BMJ Open. 2016;6:e009029. 10.1136/bmjopen-2015-009029

32. Bocanegra-Ibarias P, Garza-González E, Padilla-Orozco M, Mendoza-Olazarán S, Pérez-Alba E, Flores-Treviño S, et al. The successful containment of a hospital outbreak caused by NDM-1-producing Klebsiella pneumoniae ST307 using active surveillance. PLoS One. 2019;14:e0209609. 10.1371/journal.pone.0209609

33. Chang LWK, Buising KL, Jeremiah CJ, Cronin K, Poy Lorenzo YS, Howden BP, et al. Managing a nosocomial outbreak of carbapenem-resistant Klebsiella pneumoniae: an early Australian hospital experience. Intern Med J. 2015;45:1037–43. 10.1111/imj.12863

34. ZINC+ : Prévention, surveillance et contrôle des infections nosocomiales. Lumed. https://lumed.ca/zinc/. Accessed 5 Mar 2026

35. Nosokos : La solution des équipes en prévention des infections. Nosotech. https://nosotech.com/solutions/nosokos/. Accessed 5 Mar 2026

36. ICNET Hospital Suite : Logiciel de surveillance clinique. Baxter. https://icnetsoftware.baxter.fr/fr. Accessed 5 Mar 2026

37. sterloCare : Digitize Healthcare for Patient Experience and Compliance. OREOPS Framework. https://www.sterlocare.com/. Accessed 5 Mar 2026

38. Reynolds B, W Seeger M. Crisis and emergency risk communication as an integrative model. J Health Commun. 2005;10:43–55. 10.1080/10810730590904571

